# Extension of a SIR model for modelling the propagation of Covid-19 in several countries

**DOI:** 10.1101/2020.05.17.20104885

**Authors:** Marc Lavielle, Matthieu Faron, Jérémie H. Lefèvre, Jean-David Zeitoun

## Abstract

**Background:** Several epidemiologic models have been published to forecast the spread of the COVID-19 pandemic yet there are still uncertainties regarding their accuracy. We report the main features of the development of a novel freely accessible model intended to urgently help researchers and decision makers to predict the evolution of the pandemic in their country.

**Methods and findings:** We built a SIR-type compartmental model with additional compartments and features. We made the hypothesis that the number of contagious individuals in the population was negligible as compared to the population size. We introduced a compartment *D* corresponding to the deceased patients and a compartment *L* representing the group of individuals who will die but who will not infect anybody (due to social or medical isolation). Our model integrated a time-dependent transmission rate, whose variations can be thought to be related to the public measures taken by each country and a cosine function to incorporate a periodic weekly component linked to the way in which numbers of cases and deaths are counted and reported, which can change from day to day.

The model was able to accurately capture the different changes in the dynamics of the pandemic for nine different countries whatever the type of pandemic spread or containment measures. The model provided very accurate forecasts in the relatively short term (10 days).

**Conclusions:** In early evaluation of the performance of our model, we found a high level of accuracy between prediction and observed data, regardless of the country. The model should be used by the community to help public health decisions as we will refine it over time and further investigate its performance.

## Introduction

The coronavirus disease 19 (COVID-19) pandemic has been a disruptive event for most healthcare systems across the globe. It has led governments to take both public measures for population containment and often a resetting of healthcare organization in order to be able to absorb the peak of patients in particular for critical care. Since the inception of the outbreak, all stakeholders seemed keen to access to reliable and “living” information regarding the evolution of cases, hospitalisations and deaths in their countries as well as in others. Cross-areas comparisons were made to understand some characteristics of this novel epidemic. [1, 2] Also, being able to predict at least in the short term, the evolution of the main outcome indicators has been a relentless objective of care providers and decision makers. A single tool providing such an aid would be likely to respectively inform people and enlighten relevant stakeholders to anticipate the needed measures, in particular regarding the steering of care and social measures. We therefore developed an online model, intended to be freely accessible to the community.

The aim of this study was to report the development of the model as well as some representative data demonstrating its workability and suggesting its accuracy in studying different countries.

## Methods

We built a SIR-type compartmental model for the Covid-19 data provided by the Johns-Hopkins University (https://github.com/CSSEGISandData/COVID-19/tree/master/csse_covid_19_data/csse_covid_19_time_series). [3]

The model is a parametric model whose parameters change from country to country to reflect differences in dynamics. A classical SIR model considers 3 types of individuals: susceptible individuals (*S*), infected individuals (*I*) and recovered individuals (*R*). [4] Many extensions to this model have been proposed, in particular by adding a compartment of exposed but non-contagious individuals (*E*) to the model. [5]

Other models distinguish between symptomatic and asymptomatic patients, between patients admitted to intensive care units and patients admitted to regular hospital. [6-8] The more complex these models are, the more parameters they depend on. A classical method then consists of fixing the value of some of these parameters to values found in the literature and varying the other parameters in order to obtain simulations with dynamics that are similar to those of the data. The main objective of these approaches is not to adjust data, but rather to develop a model that is supposed to represent the reality as closely as possible. Our approach was different: our main objective was to develop an epidemiological model that fits the observed data as closely as possible, while at the same time being as simple as possible. However, we did not aim to develop an empirical model that looks like the data (e.g. using a polynomial model): the model and its parameters must have a real epidemiological interpretation, in terms of transition from one state to another or length of stay in a state. On the one hand, we made the hypothesis that the number of contagious individuals in the population was negligible as compared to the population size, which is realistic as the latest studies suggest an infection rate of around 5% in several countries. [9] This hypothesis allowed us to ignore the compartments *S* of the model. On the other hand, we had to take into account the fact that not all patients were cured but that a certain proportion may die. We therefore introduced a compartment *D* corresponding to the deceased patients.

As an examination of the data at first glance showed that there was a delay between new infections and deaths, we therefore added an additional compartment *L* in the model, which represents the group of individuals who will die (because they were infected), but who will not infect anybody (for instance due to social or medical isolation). (Figure 1)

**Figure 1.**
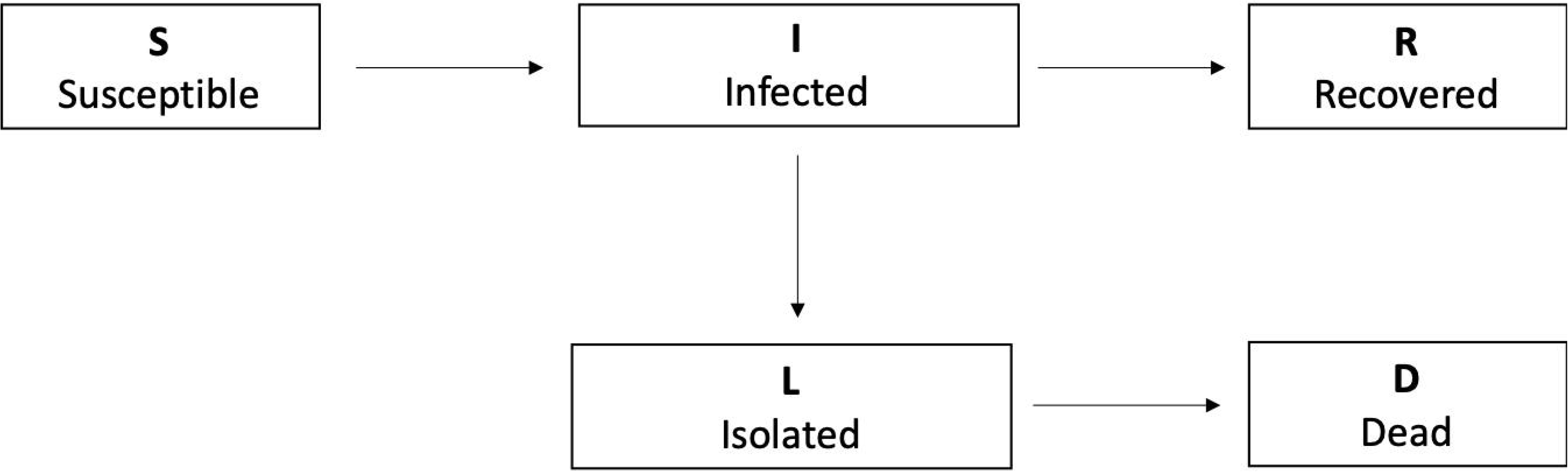
Diagram of the SIR model used.

A fundamental parameter of the model is the transmission rate, which indicates how quickly people become infected and become contagious themselves. Our model integrates a time-dependent transmission rate, whose variations can be thought to be related to the public measures taken by the country in question. One can thus imagine that the transmission rate was stable - and possibly high - before lockdown and then decreased with lockdown in countries that have adopted such a measure. A piecewise linear model was used for the transmission rate to take into account these possible variations. As the number of segments and the location of the changes in the slope are unknown, they were estimated together with the model parameters.

The data available for each country are the daily number of confirmed cases (bearing in mind that it is a number that represents an unknown fraction of the true total number of cases) and the daily number of deaths.

A periodic weekly component appears in each of these series, for many countries. This component is of course not related to the dynamics of the epidemic, which has no scientific reason to change across the days of the week, but rather to the way in which the numbers of cases and deaths are counted and reported, which can change from day to day. This component is taken into account in the observation model in the form of a cosine function. Once the model is fully defined, its parameters can be estimated for each country using precisely that country’s data.

The model, the parameter estimation algorithm, the method for model selection as well as several plotting routines have been implemented in the R package *covidix*. This package is available on Github (https://github.com/MarcLavielle/covidix). An example of how to use *covidix* can be found here: http://webpopix.org/covidixExample.html.

Note that an interactive, easy to use, web application (Shiny v1.4.0.2, 2020) also allows to visualize the data and the fitted model for several countries (http://shiny.webpopix.org/covidix/app2/). The data used in this application are updated frequently in order to be able to follow on a day-to-day basis what the model predicts for several countries.

## Results

A first version of the *covidix* package and the web application were available as of April 12^th^, with model adjustments on data from eight countries. Nine new countries were added between April 12, 2020 and May 15, 2020.

Figure 2 shows that the model is able to accurately capture the different changes in the dynamics of the pandemic *i*) for countries, such as Switzerland, which controlled the spread of the epidemic well, *ii*) for countries, such as Italy, which did not react immediately and saw the number of deaths increase very rapidly before taking containment measures which (slowly) decreased this number of deaths, *iii*) for countries, such as US, that have not taken sufficiently severe measures to significantly reduce incidences, *iv*) for countries, such as Brazil, where the pandemic has arrived more recently. Furthermore, the fits clearly highlight the weekly periodic component whenever it exists.

**Figure 2.**
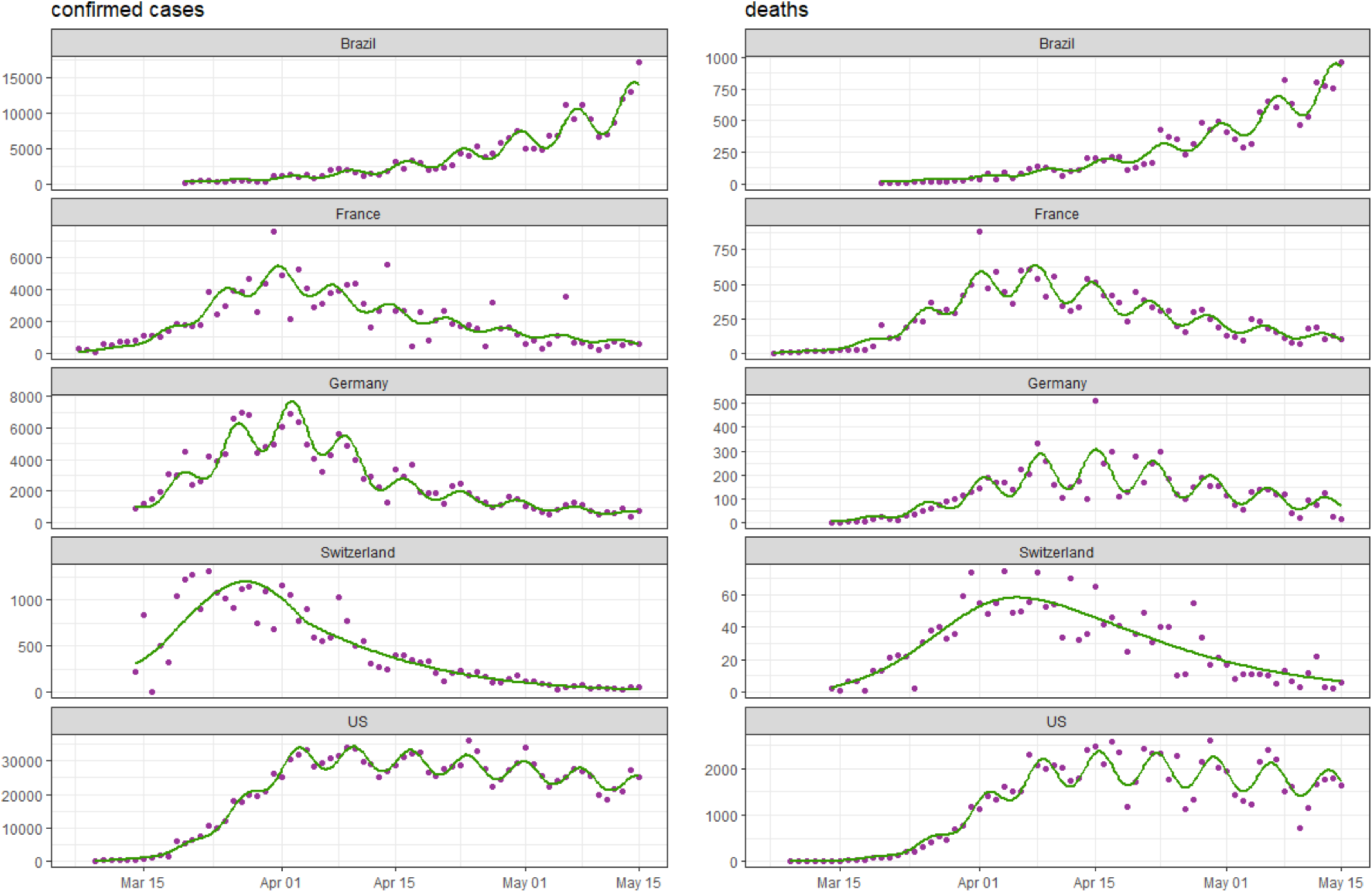
Observed and predicted number of cases for several countries (left: daily number of confirmed cases, right: daily number of reported deaths). The magenta dots are the observed data. The green curves represent the daily numbers of cases and deaths predicted by the model, including the periodic component related to how data is counted and reported throughout the week.

Figure 3 shows that the model provides very accurate forecasts in the relatively short term: the data of the last 10 days are compared to the predictions provided by a model built by ignoring these 10 data.

**Figure 3.**
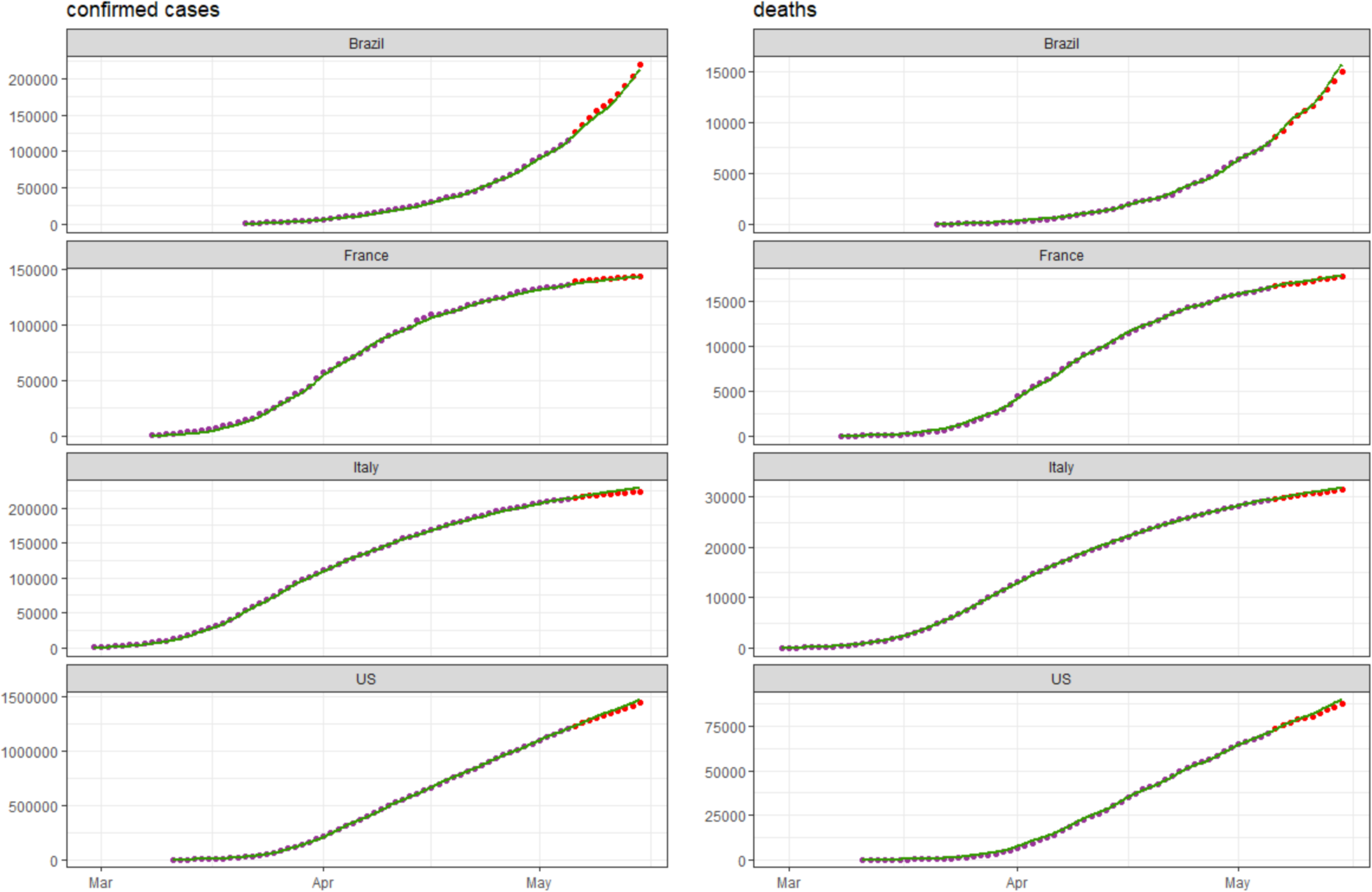
Observed and predicted number of cases for several countries (left: cumulated number of confirmed cases, right: cumulated number of reported deaths). The magenta dots are the observed data of the training dataset used for building the model; the red dots are the observed data of the validation dataset. The green curves represent the numbers of cases and deaths predicted by the model.

The model also allowed us to estimate the day of the peak of daily deaths in each country (Table 1). Moreover, the delay in days for a two- or four-folds increase or decrease in the number of deaths was assessed by the model. For instance, in Denmark, the number of deaths initially doubled every 8 days and after the peak, the number of deaths were divided by 2 or 4 every 25 days or 50 days, respectively.

**Table 1.** Maximum daily number of deaths predicted by the model. For each country, t1/4 (↑) and t1/2 (↑) are, respectively, the number of days it took to multiply the number of deaths by 4 and 2; t1/2 (↓) and t1/4 (↓) are, respectively, the number of days it took to divide the number of deaths by 2 and 4.

| Country | Increase |  | Peak of daily deaths |  | Decrease |  |
| --- | --- | --- | --- | --- | --- | --- |
|  | t1/4 (↑) | t1/2 (↑) | Date | n | t1/2 (↓) | t1/4 (↓) |
| Denmark | 12 | 8 | 4/3/2020 | 16 | 25 | 50 |
| Portugal | 19 | 15 | 4/11/2020 | 32 | 24 | 41 |
| Switzerland | 16 | 11 | 4/5/2020 | 58 | 19 | 29 |
| Netherlands | 13 | 10 | 4/4/2020 | 155 | 28 | 46 |
| Germany | 19 | 14 | 4/13/2020 | 226 | 22 | 37 |
| Belgium | 21 | 15 | 4/15/2020 | 268 | 17 | 27 |
| France | 15 | 11 | 4/5/2020 | 532 | 20 | 34 |
| Italy | 15 | 10 | 3/27/2020 | 835 | 25 | 47 |
| Spain | 16 | 12 | 4/3/2020 | 839 | 17 | 33 |
| United Kingdom | 19 | 13 | 4/13/2020 | 927 | 27 | 50 |

## Discussion

The almost unprecedented magnitude of the COVID-19 pandemic has made epidemiologic models critical not only for researchers but also for policy makers. [10] Many models have been published either in scientific journals or as open-access platforms for helping the community to describe and forecast the evolution of the pandemic outcome, yet with frequently conflicting findings.

We herein reported the development of an original model for short-term prediction of COVID-19 outcome at an aggregated level like a country, as well as updated data suggesting its very accuracy. Our model has several strengths. First, it fits the observed data, through a mechanistic modelling. Second, it appears to be validated by the data, with good short-term predictions in all tested cases. Third, it is a parsimonious model, as very few parameters are needed to describe the outcome of interest, and the prediction proved stable over time. Fourth, we implemented it as an open-access interactive tool, so as to make it operational for any relevant stakeholder for the immediate benefit of the scientific and general community in the context of the COVID-19 outbreak. Fifth, it is designed to be updated on a regular basis, ideally on a daily basis, with the possible incorporation of new countries over time. Last, it can also be seen as a monitoring tool.

We encourage researchers and policy makers to use this freely accessible model since its main purpose is to help public health decision across the globe. As the pandemic pursue its course, we will continue to enrich it with real data and will further investigate its performance.

## Data Availability

All date are published on line are available on request to the corresponding author.

